# Interictal epileptic network hubs as a biomarker for automatic localization of the epileptogenic zone: a connectivity and machine learning based analysis of stereo-EEG

**DOI:** 10.1101/2024.01.25.24301659

**Authors:** G. Susi, F. Gozzo, R. Di Giacomo, F. Panzica, D. Duran, R. Spreafico, L. Tassi, G. Varotto

## Abstract

**Objective:** The study was aimed at developing an automatic system, based on complex network analysis and machine learning, to identify interictal network-based biomarkers in patients with drug-resistant focal epilepsy and no visible anatomical lesions candidate for surgery, able to support the localization of the Epileptogenic Zone (EZ) and to further disclose properties of the interictal epileptogenic network.

**Methods:** 3 min of interictal SEEG signals, recorded in 18 patients with drug-resistant epilepsy, different EZ localization, negative MRI, were analysed. Patients were divided into seizure-free (SF) and non-seizure free (NSF) groups, according to their post-surgical outcome. After a first step of effective connectivity estimation, hubs were defined through the combination of nine graph theory-based indices of centrality. The values of centrality indices related to these hubs were used as input of an ensemble subspace discriminant classifier.

**Results:** The proposed procedure was able to automatically localise the EZ with 98% sensitivity and 59% specificity for SF patients. Moreover, our results showed a clear difference between SF and NSF patients, mainly in terms of false positive rate (i.e., the percentage of NEZ leads classified as EZ), which resulted significantly higher in NSF patients. Lastly, the centrality indexes confirmed a different role of the Propagation Zone in NSF and SF groups.

**Significance:** Results pointed out that network centrality plays a key role in interictal epileptogenic network, even in case of the absence of anatomical alterations and SEEG epileptic abnormalities, and that the combination of connectivity, graph theory, and machine learning analysis can efficiently support interictal EZ localization. These findings also suggest that poorer post-surgical prognosis can be associated with larger connectivity alteration, with wider “hubs”, and with a different involvement of the PZ, thus making this approach a promising biomarker for surgical outcome.

**Impact statement:** The correct localization of the epileptogenic zone is still an unsolved question, mainly based on visual and subjective analysis of electrophysiological recordings, and highly time-consuming due to the needing of ictal recording. This issue is even more critical in patients with negative MRI and extra-temporal EZ localization. The approach proposed in this study represents an innovative and effective tool to reveal interictal epileptogenic network abnormalities, able to support and improve the EZ presurgical identification and to capture differences between poor and good post-surgical outcome

## Introduction

Despite the great improvement in clinical and pharmacology research, epilepsy is still a diffuse neurological disease, affecting 1% of the worldwide population (Fiest et al. 2017), with approximately one third of them who exhibits seizures resistant to the antiepileptic drugs (AEDs) (Beleza 2009; Ryvlin et al. 2014). Of these patients, those affected by focal epilepsy can be considered as candidate for epilepsy surgery aimed at ablating the epileptogenic zone (EZ) (Ryvlin *et al*., 2014), which can be defined as the minimal area of brain tissue responsible for generating the recurrent seizure activity (Lüders *et al*., 2006). Various non-invasive methods, including surface video-EEG monitoring, neuropsychological tests, and brain imaging (MRI, PET, ictal SPECT), are currently applied in the attempt of identifying the EZ and the eloquent cortical areas to perform the optimal and safe resective surgical strategy (Rossi Sebastiano *et al*., 2020).

Despite the improvement in diagnostic imaging and electrophysiological techniques, invasive recordings (electrocorticography (ECoG) or stereo-EEG (SEEG)) are still required in 25–50% of the patients for the pre-surgical evaluation (Yuan *et al*., 2012; Cardinale *et al*., 2019), especially when there are discrepancies between the results of non-invasive procedures and/or magnetic resonance imaging (MRI) is unrevealing. The non-negligible rate of failure of seizure control (30-40%) one year after surgery, increasing with the duration of post-operative follow-up (Spencer and Huh, 2008; Bulacio *et al*., 2012), highlights that the issue of the correct definition of the EZ is still unsolved, and that more sophisticated diagnostic methods are required, especially for patients with non-lesional MRI.

At present, for patients submitted to stereo-EEG, the identification of the EZ is mostly based on the visual inspection of interictal and ictal invasive recordings and on the electroclinical evaluation of intracerebral electrical stimulation. However, this procedure is prone to be influenced by subjective interpretation and is considerably time consuming. The most common methodology is based on the assessment of ictal events and on the identification of fast activities identifying the seizure onset zone (SOZ) (Di Giacomo *et al*., 2019; Frauscher, 2020).

In recent years, considerable efforts have been made to develop advanced signal analysis methods to improve the EZ localization. Being focal epilepsy increasingly recognized as a “network disease”, network neuroscience offered extreme promising evidences to approach presurgical evaluation, whereas several barriers still exist and do not allow the translation of many results into clinical practice (shina et al., 2022)

While most of the studies focus on investigating network properties during ictal events, recent evidences pointed out that abnormal epileptogenic networks can be also recognized during interictal periods, making it possible to strongly reduce the length of invasive procedures and the need of recording several spontaneous seizures (see Lagarde et al., 2022 for a comprehensive review). Most of these studies agreed that interictal connectivity can be useful to localize the EZ, but they are mainly based on temporal lobe epilepsies and/or epilepsies with visible MRI lesions. To generalize these results to a more complete patient representation, including the most complicated cases of extra temporal lobe and non-lesional epilepsies, is of paramount importance to take a step forward toward real clinical application.

The aim of our study was to identify an interictal biomarker, based on the combination of complementary measures of node’s centrality (i.e., the importance of a node to act as a hub in the network), in a group of patients with different EZ localization and no visible MRI lesion, and to appraise this method as an automatic tool to support the correct identification of the EZ. With this goal, we developed an innovative approach, based on the combination of effective connectivity, graph theory and machine-learning methods, to automatically classify, in a complete blind system, SEEG leads as belonging to the EZ or not, in two groups of patients (seizure-free (SF) or not seizure free(NSF) after surgery)

Non-linear regression-index (Pijn and Lopes da Silva, 1993; Wendling *et al*., 2010) was used to quantify connectivity patterns. To extract network information, we focused on nine different graph centrality measures, suitable to quantify the capacity of a node to influence, or to be influenced by, other nodes by virtue of its connection topology (Borgatti, 2005; Fornito *et al*., 2016). With the purpose of obtaining satisfactory classification performances, we had to consider the imbalanced nature of the dataset, reflecting the increased probability that a contact belongs to the non-EZ class rather than the EZ one. To do this, we adopted a dedicated machine-learning pipeline to face the so-called class imbalance problem (Chawla, 2009; López *et al*., 2013).

The EZ as identified by clinicians with standard inspective assessment procedures and actually removed or thermocoagulated (Cossu *et al*., 2014) by surgery - was then compared to the EZ localised by our approach, hereafter called *automatically classified EZ* (ac-EZ) both for patients with successful and poor surgical outcomes.

## Materials and methods

### Patients

The study included Np=18 patients (seven women) with drug-resistant focal epilepsy and no visible anatomical lesions, who underwent SEEG at the Claudio Munari Epilepsy Surgery Centre of Niguarda Hospital (Milan, Italy). The main clinical features, the EZ localization performed by standard methods, and the surgery outcomes, are summarised in Table I. Consecutive patients were enrolled in the present study and underwent to standard diagnostic procedures as described in Rossi *et al*., 2020.

**Table 1.** Patients main clinical features.

| ID | gender | Age range at surgery (years) | Onset of epilepsy (years) | Sz/m | side | lobe | Histology | Engel Class | Follow-up /m | AEDs |
| --- | --- | --- | --- | --- | --- | --- | --- | --- | --- | --- |
| 1 | M | 21-25 | 19 | 8 | L | Fr | FCD Ia | II | 36 | ongoing |
| 2 | M | 26-30 | 19 | 10 | R | TFI | crypto | Ia | 68 | reduced |
| 3 | M | 21-25 | 4 | 10 | L | TI | crypto | Ia | 48 | stopped |
| 4 | F | 26-30 | 16 | 5 | R | T | crypto | Ia | 54 | reduced |
| 5 | F | 26-30 | 17 | 10 | R | T | crypto | Ia | 46 | stopped |
| 6 | M | 36-40 | 16 | 30 | L | Fr | no | Ib | 70 | reduced |
| 7 | M | 21-25 | 17 | 20 | R | FC | FCD Ia | III | 68 | ongoing |
| 8 | M | 36-40 | 20 | 1 | R | Fr | crypto | Ia | 42 | reduced |
| 9 | F | 36-40 | 8 | 2 | L | T | no | IV | 68 | ongoing |
| 10 | M | 11-15 | 1 | 4 | L | TPO | crypto | IV | 60 | ongoing |
| 11 | M | 26-30 | 11 | 3 | L | Fr | crypto | Ic | 65 | ongoing |
| 12 | M | 26-30 | 24 | 10 | R | T | FCD Ia | II | 59 | ongoing |
| 13 | M | 21-25 | 16 | 15 | L | TO | crypto | Ia | 36 | reduced |
| 14 | M | 41-45 | 22 | 10 | R | TO | FCD Ib | Ia | 62 | reduced |
| 15 | F | 31-35 | 4 | 5 | R | TPCF | FCD Ia | Id | 71 | ongoing |
| 16 | F | 31-35 | 12 | 20 | L | PCI | crypto | IV | 62 | ongoing |
| 17 | F | 31-35 | 1 | 8 | R | T | HS | II | 70 | reduced |
| 18 | F | 36-40 | 9 | 30 | L | TC | crypto | III | 53 | ongoing |

The patients’ mean age at seizure onset was 13.1 ± 7.2 years, and the mean duration of epilepsy was 17 ± 9.1 years. They had no obvious risk factor for epilepsy. All had a negative MRI. One patient had a neuropathological diagnosis of Hippocampal Sclerosis (HS), not identified by pre-operative neuroradiological examination and not associated with seizures suggesting temporo-mesial origin.

The placement of intracerebral electrodes was defined according to the data derived by non-invasive anatomo-electroclinical procedures.

The results of mathematical analysis, performed after surgery, did not interfere with the surgical planning. At the end of the invasive recordings, a SEEG guided thermocoagulation (THC) (Cossu *et al*., 2017) was performed in 11 patients, but only one (n. 7) of them became seizure and did not undergo surgery. Patient 10, was did not undergo to resective surgery due to the involvement of the language areas in the identified EZ. Histopathological diagnosis was thus performed on surgical specimens from 16/18 patients.

The surgical outcome was assessed after at least three years of follow-up after surgery (or THC), and classified according to Engel’s classification (Engel, 1993). The mean follow-up was 58±12 months. According to their outcome, patients were divided into two classes: *seizure-free* (i.e., SF group – belonging to classes Ia-b-c-d, *N_p,SF_* = 10) and *non-seizure free* (i.e., NSF group - class II-IV, *N_p,NSF_* = 8).

### SEEG recordings

Multi-lead platinum-iridium electrodes (Dixi, Besançon, France; 5–18 contacts each; diameter 0.8 mm; 1.5 mm long and 2 mm apart) were implanted under general anaesthesia after stereo-arteriography using three-dimensional MRI imported into a computer-assisted neuro-navigational module to localise the blood vessels and guide electrode trajectory. SEEG was tailored to individual anatomic and electroclinical characteristics following the procedure described in Cardinale *et al*., 2019.

The SEEGs were recorded using a common reference electrode (Nikon-Kohden system; 192-channels; sampling rate 1 kHz) under video and clinical control over 5-20 days. After the recordings and the low- and high-frequency stimulation had been made, and any THC performed, the electrodes were removed. Two trained neurologists evaluated the SEEG recordings to define the EZ and plan the surgical approach and resection examined the SEEGs.

Although SEEG ictal recordings were available for all the patients, we considered for this study only the interictal signals. The selected recordings were acquired during awake resting state conditions, at least one day after electrode placement and one hour far from any ictal event.

### Clinical identification of the EZ and PZ

The Epileptogenic Zone was defined as the cortical area from which the ictal discharges originated. The clinical modifications associated with the intracerebral electrical stimulations and the neurophysiological mapping were integrated in the identification of the brain area(s) to be surgically excised. The early propagation zone (PZ) was defined as the region in which seizure’s activity were visible within 5 seconds after seizure onset. Post-resection MRI was the used to identify areas that were removed or thermocoagulated.

### SEEG connectivity analysis

A specific toolbox for SEEG data visualisation and selection was developed and integrated into our custom-made software package in the Matlab environment (MathWorks Inc., Natick, MA, U.S.A.). Signals were analysed with bipolar montage, and derivation with artifacts were excluded from the analysis; the mean number of resulting SEEG leads (*N_L,p_*) were *N_L_*= 72 ± 8. For each patient, 3 minutes of continuous interictal signal were selected and divided into *N_E_* = 36, five s length, non-overlapping epochs (Fig.1A). The effective connectivity pattern was estimated by means of a bivariate non-linear method, the *non-linear regression index* (*h_2_*) (Wendling *et al*., 2010), thus obtaining 36 time-varying adjacency matrices (Fig.1B, top). A broad frequency band [1-80 Hz] was used for the analysis, since the *h_2_* is best suited to be computed considering the broadband signal (Lagarde et al., 2022) In order to test whether distances between the areas could influence classification performances, a second set of adjacency matrices was calculated after normalising the connectivity values by the Euclidean distances, for each couple of leads. All the following analysis was performed on both adjacency matrices, with and without distance normalisation. From now on, DN (distance normalization) will refer to connectivity values normalised by intercontact distance. Moreover, for both SF and NSF patients, linear correlation between connectivity and distance for each couple of leads was calculated, considering all leads together as well as separated EZ, PZ and nEZ ROIs.

**Fig. 1.**
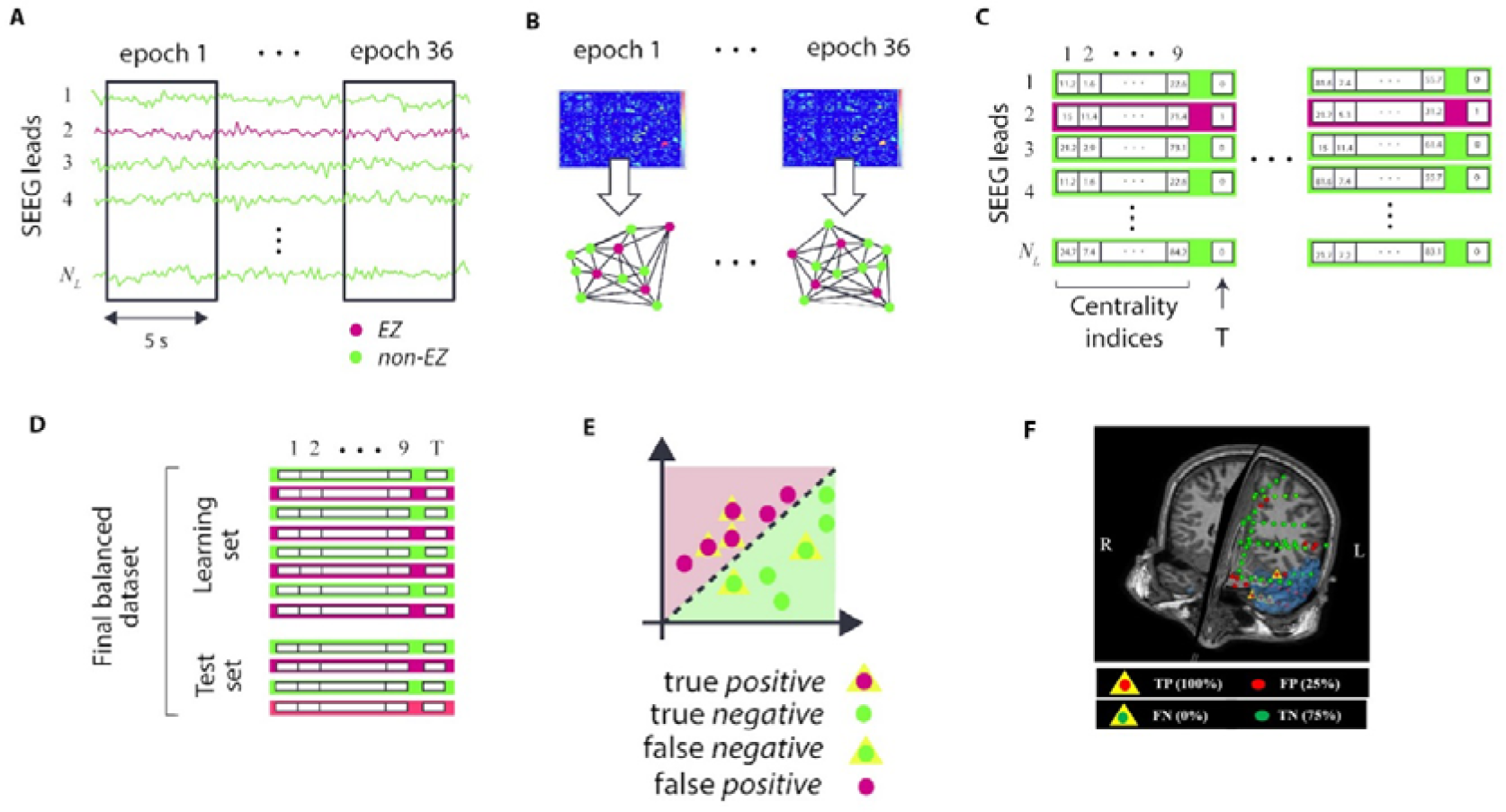
Schematic representation of the processing blocks of the applied method: (a) for each patient, selection of SEEG time-series and partition into 36 non-overlapping epochs. (b) Estimation, for each epoch, of the effective connectivity matrices, to generate the related graphs, where each vertex represents a SEEG lead. c) Calculation of the 9 centrality indexes for each vertex of the graph, and arrangement in sets of strings composed of the feature values (graph indexes) and the pertaining class of the corresponding contact (unbalanced dataset). (d) Combination of data of the whole set of patients, rearrangement in the ‘vertical’ data-set, and training data-set rebalancing with ADOMS approach. (e) Classification using the four methods selected as best learners. (f) Visualization on the surgical 3D scene of the final classification of the SEGG leads to automatic-classified EZ (ac-EZ) or automatically classified non-EZ (ac-non-EZ), for a representative patient.

### Graph analysis

After the generation of the adjacency matrices, the corresponding graphs were built for each patient (Fig.1B, bottom). In order to detect only significant connections, a threshold was defined considering, for each epoch, the minimum degree in order to have a connected graph (i.e. graph in which there is a path between each pair of vertices, to avoid isolated nodes in the network). The maximum value among all epochs was then selected as final threshold, in order to have a fixed global degree among all patients

Nine graph theory-based indexes, focusing on different centrality’s network properties (Oldham *et al*., 2019), have been calculated, and used as features of the classifier: *outdegree, indegree, oustrength, instrenght, betweenness, outcloseness, in closeness, pagerank* and *eigenvector centrality*.

Each different centrality metrics define the importance of a node from a different and complementary perspective and, therefore, for the same network, can identify different nodes as being the “most central” ones.

See Supplementary Table I, for list and description of the basic properties of these indexes.

### Machine Learning-based classification of EZ

We used the approach of supervised classification to automatically detect whether time series of single SEEG contacts correspond or not to the EZ.

The dataset to be classified was characterised by imbalanced class distribution (over the nodes, on average, only 15% pertains to the EZ and 85% to the non-EZ), and this could result in a significant loss of classification performance if faced with standard machine learning approaches (Chawla, 2009; Lopez *et al*., 2013). For this reason, we applied the ADASYN rebalancing approach (Tang and Chen, 2008) to generate synthetic examples of the minority class (EZ), by selecting *k-nearest neighbours* and using them to synthesise new instances by PCA. Then, the obtained rebalanced dataset was classified through an ensemble subspace discriminant classifier (Kuncheva, 2004). The selection of the optimal rebalancing and classification methods was done after testing and comparing several different approaches, as described in Varotto et al. 2021. The classifier was fed with nine features (i.e., the nine centrality indices) and a target value (the class corresponding to each set of centrality values – i.e., SEEG leads belonging to EZ or non-EZ) (Fig. 1C). The latter has been defined by considering the intersection between the group of leads labelled as EZ by clinician through the pre-surgical evaluation, and the final resected zone. Cross validation was performed through the leave-one-patient-out (LOPO) approach: we generated 18 different *group datasets*, each one using one different patient as test, and the whole set of trials from the remaining 17 patients for the training. This approach allowed us to dispose of a greater number of subjects for the training and to obtain classification performances referred to a specific patient (the test set is composed of only one patient each time).

To assess the classification performances, the *area under the curve* (AUC), the *true positive rate* (TPr, or *sensitivity*) and *true negative rate* (TNr, or *specificity*) were considered as metrics (López *et al*., 2013).

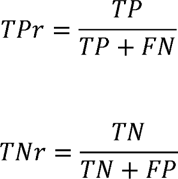

The machine learning analysis was performed by combining the software *Keel* (Alcalá-Fdez *et al*., 2009), for the data rebalancing, and the *Matlab classification learner* toolbox, for the classification.

### Statistical analysis

All the statistical comparisons between SF and NSF groups were performed by two-tailed non-parametric Mann-Whitney U test and Wilcoxon test using SPSS software (IBM Corp. Released 2019. IBM SPSS Statistics for Windows, Version 26.0. Armonk, NY: IBM Corp). The alpha level for statistical significance was set to 0.05. A false discovery rate (FDR) approach was applied in order to correct for multiple comparisons. All the reported *p-values* refer to the corrected ones.

## Results

### Centrality Indexes

For each of the nine-centrality indexes, statistical differences between EZ and nEZ were calculated, in both SF and NSF groups. Results enhanced a clear different pattern between the two patient groups, with the first one showing a significantly higher centrality in EZ than nEZ in 8 out 9 of the centrality indexes (being the BC the only one not significant). On the contrary, for the NSF group, 7 out of the 9 indexes did not reveal significant differences between EZ and NEZ leads. (Fig. 2a)

**Fig. 2.**
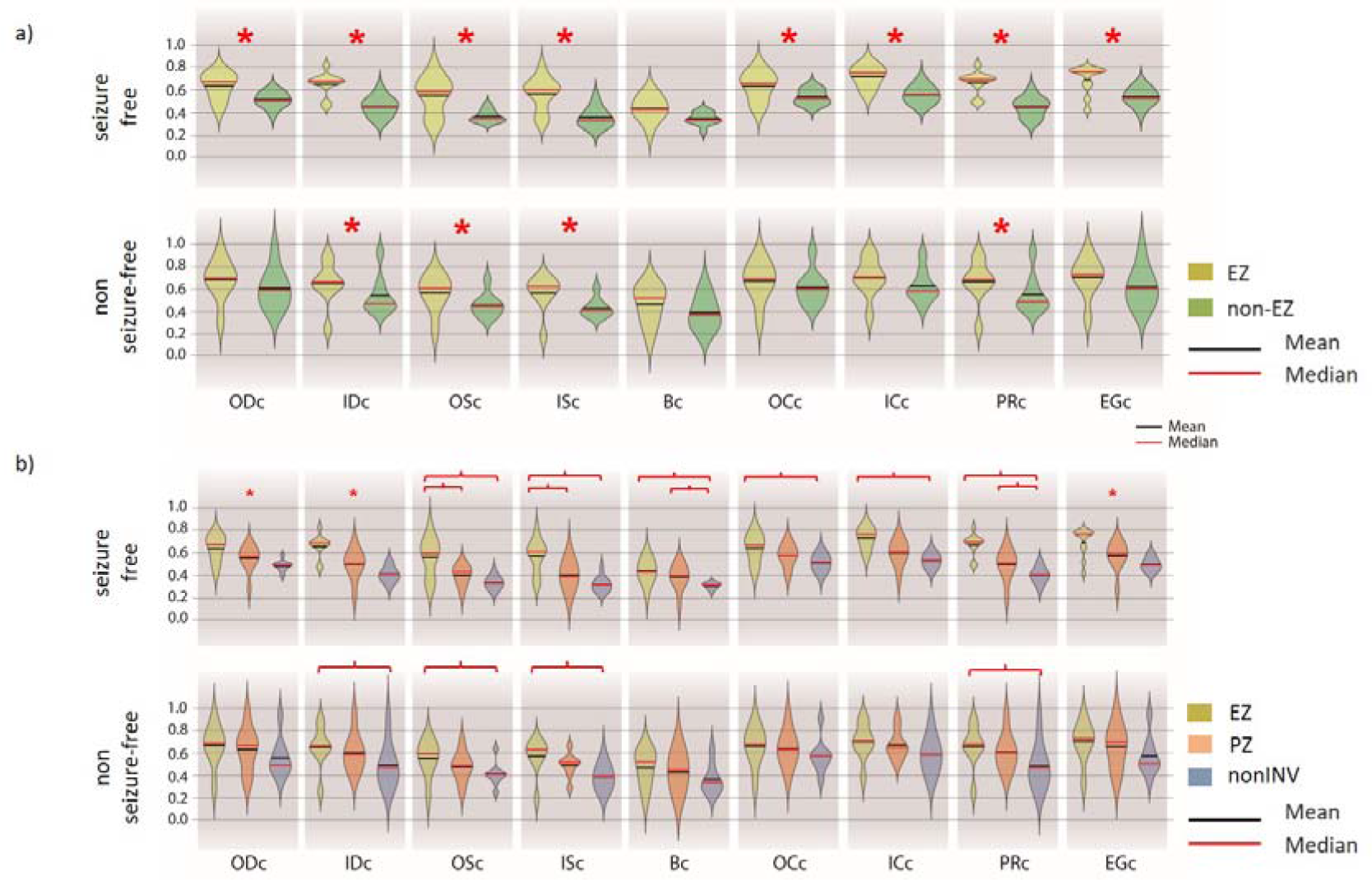
Violin plots representing the nine centrality indexes, among different ROIs. In Fig 2a, the difference between EZ and nEZ was considered. The red asterisk indicates a significant difference, after FDR correction. In Fig 2b the nEZ was further divided into propagation zone (PZ) and not-involved zone (nonINV). Significant differences between pairs or regions are represented with red brackets. A red asterisk indicates a significant difference among all the three regions.

In Fig 2b the same values are represented, but splitting the nEZ zone into two sub-regions: *propagation zone* and *not involved zone*. For the SF patients, all 9 indexes revealed a first-level significant difference. EZ centrality was significantly higher than nEZ for all the indexes, and higher than centrality in PZ for ODc IDc OSc, ISc and EGc. Interestingly, in the NSF group none of the centrality indexes reached a significant difference between EZ and PZ. The detailed breakdown of classification results by subject is reported in Table 2.

**Table 2.** Detailed breakdown of classification results by subject, in terms of TPr (Sensitivity) and TNr (Specificity), obtained from raw connectivity values.

|  | SF |  |  | NSF |  |
| --- | --- | --- | --- | --- | --- |
| Id Pt | TP rate | TN rate | Id Pt | TP rate | TN rate |
| 2 | 1,00 | 0,62 | 1 | 0,89 | 0,14 |
| 3 | 1,00 | 0,73 | 7 | 1,00 | 0,00 |
| 4 | 1,00 | 0,34 | 9 | 0,00 | 0,47 |
| 5 | 1,00 | 0,65 | 10 | 0,83 | 0,73 |
| 6 | 1,00 | 0,59 | 12 | 0,78 | 0,58 |
| 8 | 1,00 | 0,49 | 16 | 0,67 | 0,75 |
| 11 | 1,00 | 0,54 | 17 | 0,00 | 1,00 |
| 13 | 0,92 | 0,68 | 18 | 0,93 | 0,60 |
| 14 | 1,00 | 0,57 |  |  |  |
| 15 | 0,71 | 0,60 |  |  |  |
| Mean | 0,96 | 0,58 | Mean | 0,64 | 0,53 |
| StDev | 0,09 | 0,11 | StDev | 0,41 | 0,33 |

### EZ classification

AUC obtained from the SF group was higher, even if with no significant difference, than the NSF group. (Mean *AUC_SF_* = 0.78 ± 0.06; Mean *AUC_NSF_* = 0.64 ± 0.18, p-value < 0,165) (Supplementary Fig 1).

TPr and TNr individual values, for all the Sf and nSf patients, are shown in Table 2. Fig 3 a represents the average percentage of TPr, TNr, FPr, FNr for the two groups of patients, with the raw connectivity data (not taking into account the distance).

**Fig. 3.**
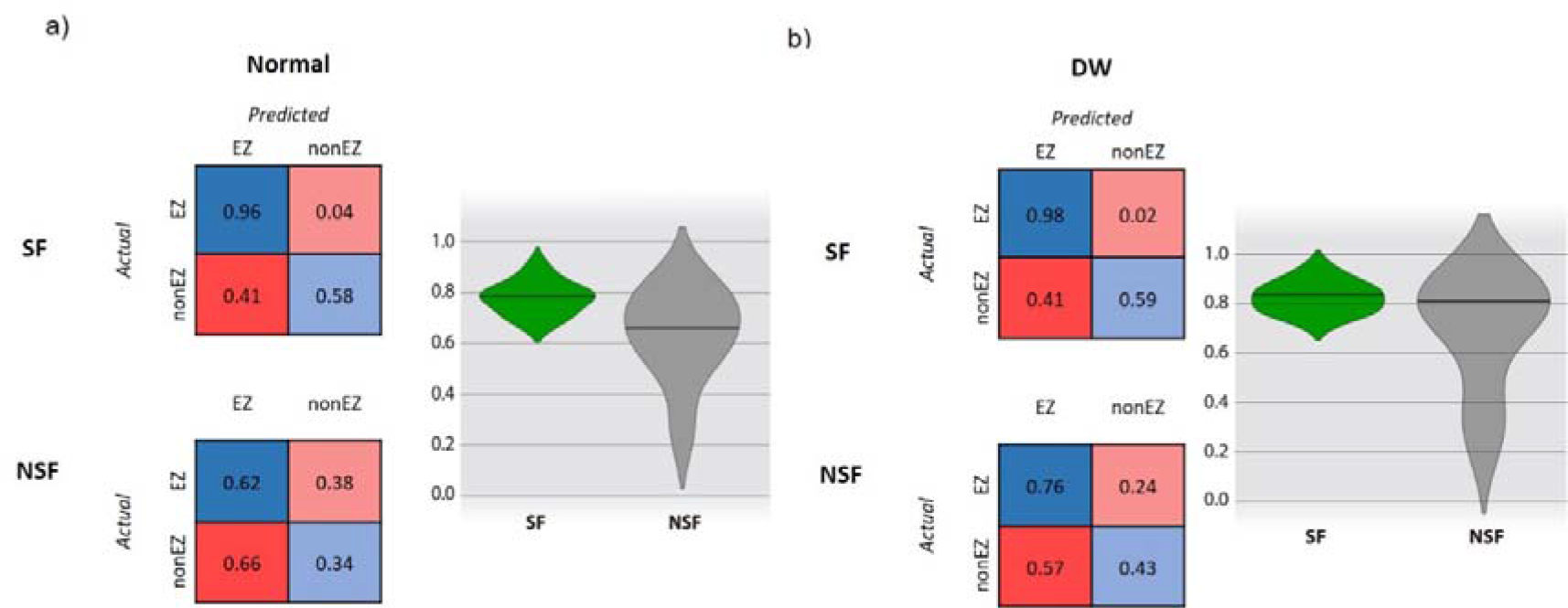
Classification results by class (ratios), in terms of TPr (Sensitivity) and TNr (Specificity). TPr (blue), TNr (light blue), FPr (red), FNr (light red), and related violin plots (with means). Classification performances are reported for raw connectivity data (a) and weighted by Euclidean distances (b).

The results enhanced a very high TPr for the SF patients, with the classifiers able to correctly detect the EZ leads with a sensitivity range of 96±9%, while a lower and much more variable values were found among subjects for the NSF group, with a sensitivity range of 64±41% (p_value < 0,009). In terms of TNr, the results showed a lower specificity of the classifiers, with no significant differences between the two groups. (Mean *TNr_SF_*: 58±11%; Mean *TNr_NSF_*: 53±33%; p_value < 1).

Finally, in order to evaluate the effect of the propagation zone in the classification, namely in the identification of false positive EZ regions, we calculate the percentage of PZ leads that were classified as EZ from our method (false positive EZ). We found similar results in SF and NSF patients, with 30% of false positive EZ corresponding to the PZ regions (Supplementary Fig. 2).

Fig. 4 shows the final classification on the 3D scene for two representative patients, one SF (patient 3) and one NSF (patient 12).

**Fig. 4.**
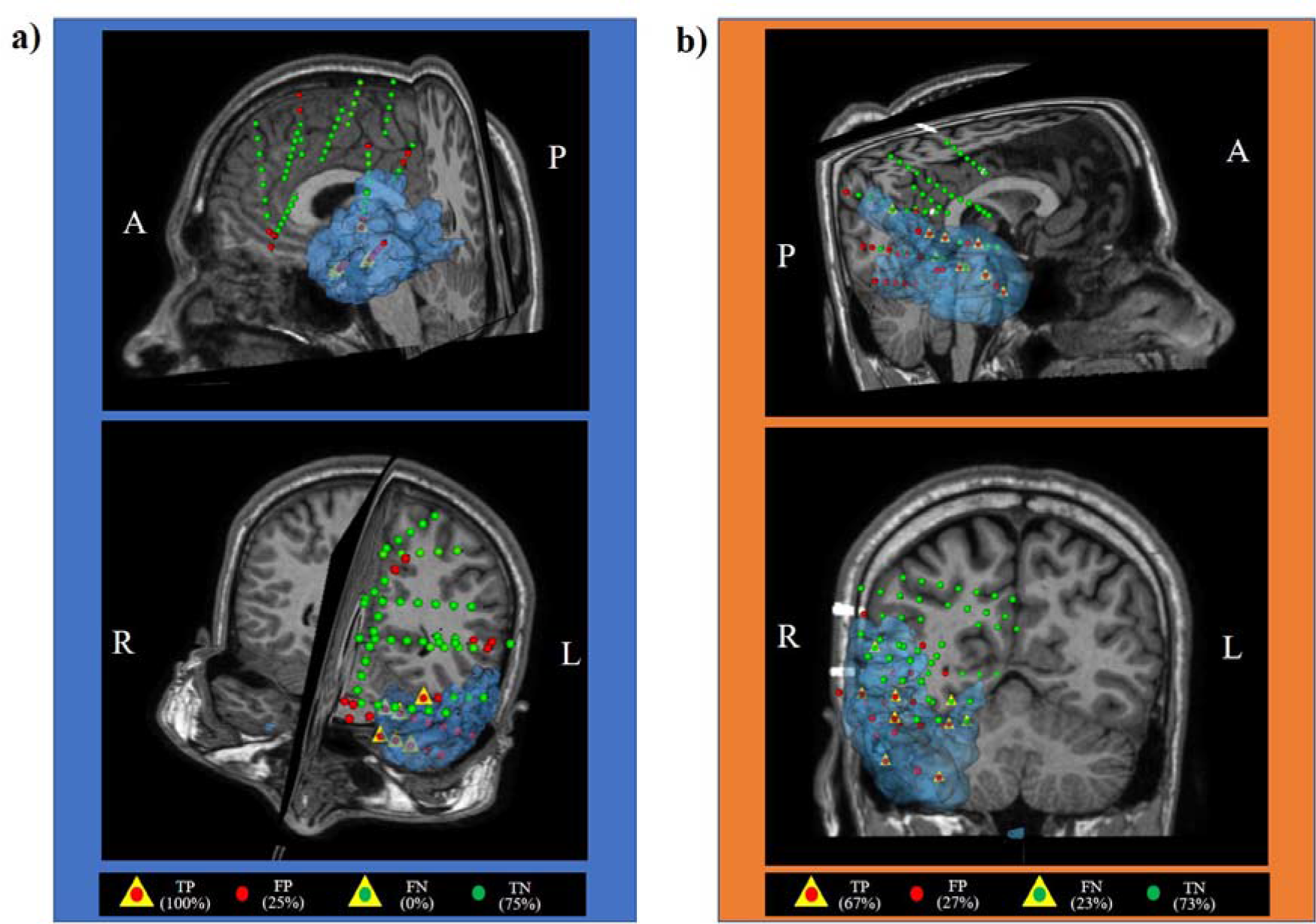
Visualization of the final classification of the electrode as ac-EZ or ac-nonEZ for one representative SF patient (pt. 3) (a) and one NSF patient (pt12) (b) and on his surgical 3D scene. The blue area indicates the final resected zone. A: anterior; L: left; P: posterior; R: right.

### Inter-patient variability

Classification performances show clear differences between SF and NSF group in terms of inter-patients’ variability, with the latter group characterised by a much higher variability, especially for the TPr values, as resumed by the mean between the Interquartile Range (IQR), a measure of data variability (TPr: IQR_SF_ = 0 and IQR_NSF_ = 0,40; FPr: IQR_SF_ = 0,10 and IQR_NSF_ = 0,35). See Fig.5 for the corresponding box plot

**Fig. 5.**
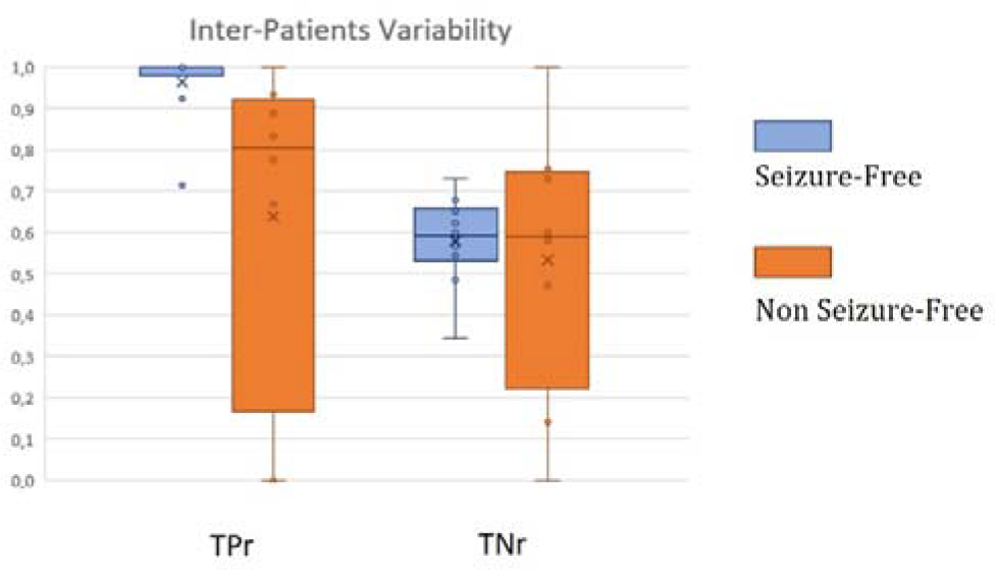
Box plot showing Inter-subject variability of TPr (a) and FPr (b), showing mean, median, 25%, and 75% percentile, and values distribution of classification performances.

### Effect of distance in classification

Correlation between the *h_2_* values and distance show a decrease in connectivity values with the increase of intercontact distance. A similar trend was found for both SF and NSF patients for all regions except for EZ, where a stronger correlation characterised the SF group with respect to the NSF (Fig. 6).

**Fig. 6.**
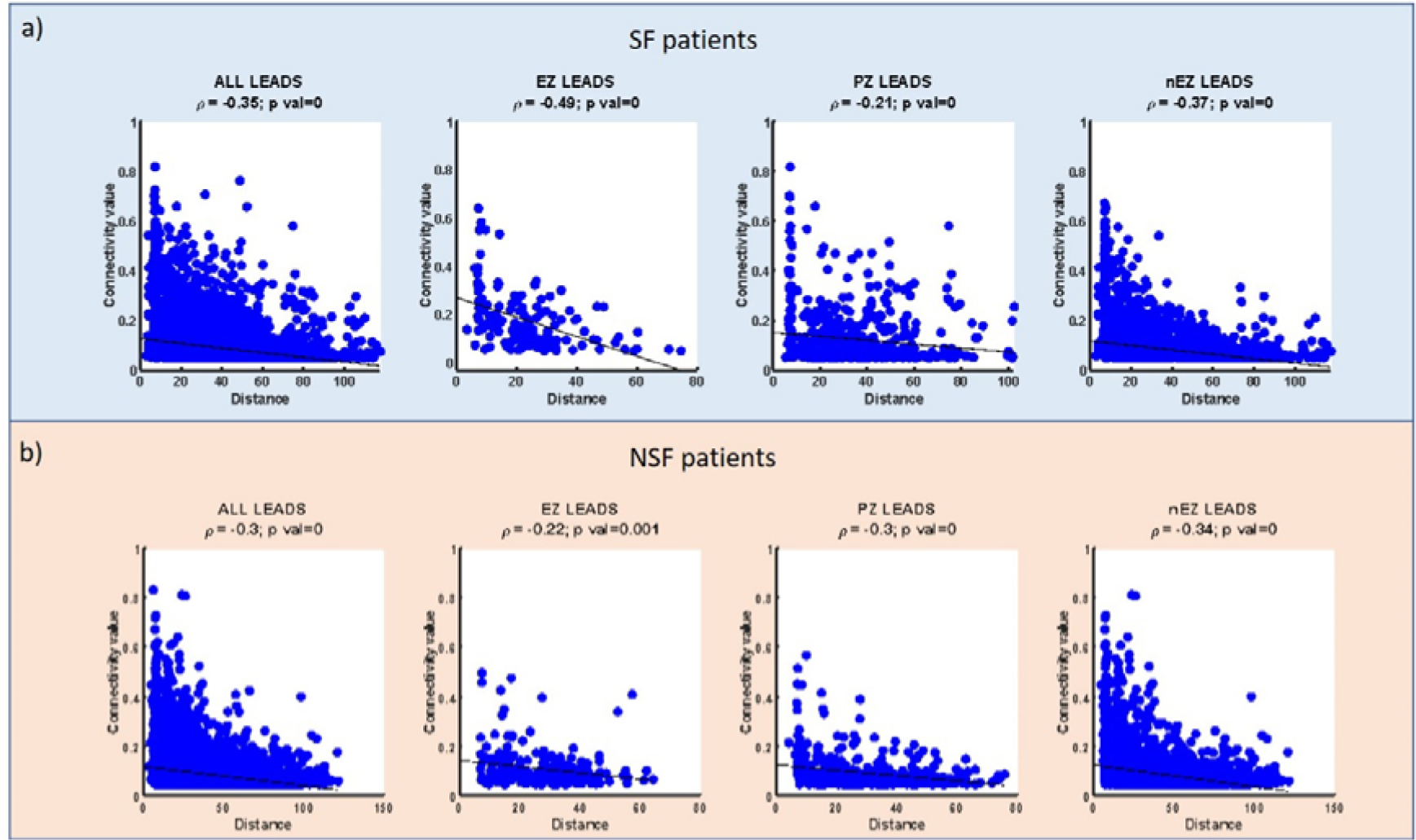
Correlation between connectivity values and Euclidean distances for each pair of sEEG lead, for SF (a) and NSF 8b) patients. The correlation was estimated considering all the leads, as well as the EZ, PZ, and NEZ regions independenty

In order to evaluate the effect of intercontact distance for the correct localization of the EZ, the whole classification procedure was repeated by considering as input of the analysis the connectivity values normalised by the distances between each pair of SEEG leads.

The resulting AUC values are comparable to those obtained without considering the distances (Mean *AUC_SF_DN_* = 0.84 ± 0.1; Mean *AUC_NSF_DN_* = 0.69 ± 0.23, p-value < 0,173). (Supplementary Fig. 1)

With respect to TPr and TNr, the distance affected in a slightly different way the results from the SF and NSF group. In the first one, the introduction of distance weights did not change classification performances (TPr*_SF_* 98±5%; TNr*_SF_* 58±17%). On the contrary, for the NSF group, the introduction of the distance weights resulted in an improvement of TPr and a decrease of TNr with respect to the classification based on raw connectivity (TPr_NSF_ 83±28%, TNr_NSF_ 44±25%), whereas these differences did not reach statistical significance (Table 3).

**Table 3.** Detailed breakdown of classification results by subject, in terms of TPr (Sensitivity) and TNr (Specificity), obtained from connectivity values weighted by the Euclidean distance.

|  | SF |  |  | NSF |  |
| --- | --- | --- | --- | --- | --- |
| Id Pt | TP rate | TN rate | Id Pt | TP rate | TN rate |
| 2 | 1,00 | 0,52 | 1 | 0,93 | 0,58 |
| 3 | 1,00 | 0,82 | 7 | 1,00 | 0,11 |
| 4 | 1,00 | 0,22 | 9 | 0,20 | 0,47 |
| 5 | 1,00 | 0,76 | 10 | 1,00 | 0,54 |
| 6 | 1,00 | 0,69 | 12 | 0,89 | 0,61 |
| 8 | 0,86 | 0,62 | 16 | 0,67 | 0,62 |
| 11 | 1,00 | 0,47 | 17 | 1,00 | 0,00 |
| 13 | 1,00 | 0,51 | 18 | 0,93 | 0,60 |
| 14 | 0,92 | 0,69 |  |  |  |
| 15 | 1,00 | 0,55 |  |  |  |
| Mean | 0,98 | 0,59 | Mean | 0,83 | 0,44 |
| StDev | 0,05 | 0,17 | StDev | 0,28 | 0,25 |

## Discussion

In this study, we developed an innovative approach to automatically classify the SEEG data as belonging or not to the EZ, based on the analysis of segments of interictal activity. This approach is based on the combination of effective connectivity, graph theory-based centrality indexes, and machine learning methods. We selected 18 patients with focal epilepsies, different EZ location and extension, negative MRI lesion, and we compared 10 SF versus 8 NSF patients at the evaluation performed at least three years after surgery.

The main findings of our study indicate that: (i) interictal period is characterised by abnormal connectivity pattern suitable to automatically identify the EZ, even in absence of anatomic alterations; (ii) centrality (i.e. the importance of a node to act as hub in the network), plays a key role in the epileptogenic network and in the EZ localization; (iii) poor post-surgical outcome appears to be associated with extensive connectivity alteration and with more extended “hubs” involving regions exceeding the EZ identified by standard procedures; (iv) interictal network centrality could be a promising biomarker to assess risks of post-surgical negative outcome. The pipeline we developed is fully automatic and can be easily applied to data with minimal preparation, offering a promising approach to be tested by other groups for larger cohort validation study.

### Interictal EZ localization

The chance to use interictal recordings to support the EZ localization could be extremely important in order to reduce high time consuming and cost of invasive monitoring, which normally requires many days to be able to record enough seizures. Several studies reported evidence about the ability of connectivity metrics to identify EZ abnormal network alterations during interictal recordings (Varotto et al, 2012; Van Diessen *et al*., 2013; Goodale et al., 2020; Narasimhan et al 2020; Jang et al 2022). Nevertheless, all these studies focused on specific patient groups, mainly affected by temporal lobe epilepsies and with MRI-visible lesions, although extra-temporal epilepsies and negative MRI patients represent a frequent, problematic and complex issue for epilepsy surgery. In few more recent studies, larger cohorts of patients, spanning more epilepsy subtypes and EZ localization, identify the correlation between interictal connectivity patterns, the EZ and with the post-surgical outcome (Lagarde *et al*., 2018, Paulo et al, 2022). At present, however, there is no assessed method to define the EZ only on the basis of interictal neurophysiological data.

It is important to emphasize that while most of the cited studies devoted to the identification of EZ using interictal signals were based on the presence of interictal spikes and discharges, our study was conversely based on “resting” interictal epochs, free from epileptic abnormalities. Thus, our results further suggest that resting state network connectivity may reveal a persistent reorganisation occurring in focal epilepsy (Englot *et al*., 2016) and could represent both the cause and the consequence of recurrent seizures.

This condition is also suggested by results obtained in animal models, revealing enhanced network connectivity even without change of excitability involving individual neurons (Traub *et al*., 2001; Blumenfeld *et al*., 2007). Changes in synaptic coupling (Khalilov *et al*., 2003) can in fact favour a “permissive” network capable of facilitating in turn seizure recurrence and discharge propagation (Wendling *et al*., 2012; Proix *et al*., 2014).

### Wider network alteration in focal epilepsy

Reasons for the surgical failure are often unclear, but they are likely related to imperfect identification and/or resection of the EZ. At present, epilepsy can be assumed as a network disease, leading to widespread structural and functional changes, frequently associated with significant neuropsychological impairments (Ung *et al*., 2017) suggesting a complex dysfunctional condition. Some studies reported spikes located far from the supposed EZ in patients with focal epilepsy (Moseley et al., 2012), moreover patients who underwent surgery can later present seizures originating from dissimilar regions from the original EZ (Vaugier *et al*., 2018). All these evidences confirm that, in focal epilepsy, seizures could arise from complex epileptogenic networks rather than from focal sources (Burman and Parrish, 2018; Englot *et al*., 2020). Our results confirmed an abnormal interictal network involving regions exceeding the EZ zone recognized by standard methods. These findings are in line with the evidence obtained from (Lagarde *et al*., 2018) who studied the interictal connectivity pattern by dividing the brain regions in EZ, propagation zone and not involved zone; the authors found a similarly high connectivity pattern in the EZ and the propagation zone, compared with the not involved zone. In a previous study (Varotto *et al*., 2012), we found that focal cortical dysplasia type II exhibit increased connectivity not only within the anatomical lesion, but also in other cortical areas, which appeared to be secondarily involved in seizure propagation, and that these areas are crucial to trigger the seizure onset. Our *false positive regions* (e.g. zone classified as EZ by our tool but not by the clinicians) however do not reveal a correspondence with the PZ (only 30% of PZ leads corresponded to our false positive regions), suggesting that the wider interictal hubs we detect are capturing other regions - and some more complex mechanisms - than these PZ. The relationship between EZ, PZ (including an agreement about definition of OZ) and interictal network is still an open question, that need to be proper investigated.

Concerning the NSF group, our main finding was a false positive rate significantly higher than in the SF patients. Some previous studies already suggested a correlation between poorer post-surgical outcome and wider areas of high synchrony (Antony et al 2013; Lagarde *et al*., 2018), indicating that connectivity metrics can improve prediction of the post-surgical outcome (Bonilha et al 2015; He *et al*., 2017). Our results confirm that negative post-surgical outcome is associated to larger defective interictal networks, with NSF patients having wider “hubs” than the SF ones, which could account for an incomplete resection of the EZ or a more complex and extended altered network not recognized by standard signal evaluation and able to allow seizures recurrence. Overall, this confirms the need to incorporate brain network analysis in epilepsy surgery, as a crucial approach to better target surgical strategies.

### Centrality: interictal “hubness” of the EZ

Several studies already investigated the role of central nodes, or hubs, and their relation to epileptogenic network, although most of the collected data focused on ictal activity. (Adkinson *et al*., 2019) applied several centrality indexes to early ictal epochs and found the highest centrality of gamma band activity in the seizure onset zone. Burns *et al*. (2014), when applying the eigenvector centrality time course before, during, and after the seizure end, found a significant relationship between centrality’s time dynamic and the location of EZ. Moreover, ictal eigenvector centrality was found to be a possible biomarker for postsurgical outcome prediction (Grobelny et al 2018).

Centrality can be measured with different indexes, thus providing complementary information on the importance of a node in the network (Oldham *et al*., 2019). To our knowledge, this is the first study combining and comparing a wide spectrum of centrality indexes. Although some of these indexes could provide redundant information in certain conditions, in our procedure we found that the best approach to localise the EZ was obtained by combining all of them, so no feature dimension reduction was applied. This means that, even if node centrality characterises the EZ in general terms, each patient could be better characterised by specific centrality property, therefore leading to a patient-specific biomarker. In a study of interictal EEG data (Marino *et al*., 2019), showed that epileptic network is often patient specific, even comparing subjects with apparently similar seizure onset locations. Our findings confirmed this evidence, but also showed a promising approach to combine these patient-specific centrality properties, in order to have a biomarker, suitable to work at group level and that wider ‘hubness’ can be a potential biomarker of poor surgical outcome (He *et al*., 2017).

### Machine learning and epilepsy

Machine learning methods have been applied increasingly in the field of epilepsy, whereas most of the applications were settled to seizure detection or imaging analysis for lateralization of temporal lobe epilepsy (Abbasi and Goldenholz 2019). Some recent studies applied machine learning methods also to evaluate connectivity measures from intracranial EEG to predict surgical outcomes, showing high sensitivity in predicting Engel I outcomes (Antony *et al*., 2013; Tomlinson *et al*., 2017), and to identify epileptogenic regions from interictal and ictal data (Elahian *et al*., 2017; Khambhati *et al*., 2017).

Starting from features derived from connectivity and graph theory, in our work it has been crucial to explore combinations of dataset resampling techniques and machine learning methodologies to face the intrinsic class imbalance character of the problem, inherited from the fact that the majority of the contacts pertain to the non-EZ. Interestingly, among the different classifiers we tested, the best performances come from the ensemble subspace discriminant analysis. The ensemble approach represents one of the main directions in machine learning theory and consists in the combination of different classifiers in order to improve their final performance. In the family of ensemble learners, we applied the random subspace analysis (Sun and Zhang, 2007), which is a combination of different subsets of the classifier’s features (in our case the different centrality measures). This confirms that, as discussed above, each patient can be better characterised by a specific associate combination of centrality properties, and that the exploration of others ensemble approaches can be a promising future direction in this field.

Moreover, advanced machine learning approaches applied to EZ localization can be a promising tool since they will allow combining and testing several signal properties with different indexes, by adopting a multi-marker approach. Only one paper recently showed how combining univariate and bivariate interictal signal features, classifying them with support vector machine could strongly improve EZ localization (Cimbalnik *et al*., 2019). Our results indicate that centrality indexes are able to correctly localising the “true” EZ, with no false negative error, while the false positive errors could be reduced by the multi-marker approach, by combining centrality with other possible EZ interictal markers.

## Conclusions

Localization of the EZ is still a challenging issue, especially in patients with no lesions visible at MRI, and advanced signal processing methods could be a useful tool to support clinicians and to reduce the rate of surgery failure. As well, the possibility to localise the EZ from interictal signal epochs can strongly reduce the recording time needed to detect seizures.

Overall, our results show that interictal signals may contain enough information to localise the EZ, and that higher centrality ‘hubness’ is a property that clearly characterised these regions, confirming that seizures could arise from complex epileptogenic networks rather than from focal sources. By combining connectivity, graph theory, and machine learning methods we developed and automatic procedure able to detect the EZ, as identified by the current gold-standard clinical practice, and to be appropriate in a heterogeneous group of patients. The approach was designed to work on a group level, but it showed to be effective when tested also at a single-patient level, a condition that is essential to tailor the surgical strategy according to specific patient characteristics.

Moreover, machine learning is a suitable tool to combine different signal properties and metrics enabling to develop multi-marker approaches, which can be a useful future perspective.

Next steps will be the application of these methods to a bigger group of patients, and the addition to this classification procedure of other possible EZ interictal markers, able to improve the specificity of the method.

## Abbreviations

EZ: epileptogenic zone
NEZ: non-epileptogenic zone
TPr: true positive rate
FPr: false positive rate
SF: seizure free
NSF: non-seizure free
SEEG: stereo-EEG
THC: thermocoagulation.

## Author Contribution

G.V, F.P., S.F, L.T. and R.S. launched the idea of this experiment. G.V. conceptualized and developed the methodological procedure. G.V. and G.S. performed the analysis. F.P and G.S. contribute to methodological improvement. L.T. and F.G were involved in data acquisition and clinical EZ localization. G.V., F.P., L.T., S.F. and R.S. were involved in interpretation of the data. G.V. and G.S wrote the main manuscript and generated the figures. All authors collaborated in manuscript writing and correction.

## Acknowledgements and funding

The authors disclosed receipt of the following financial support for the research, authorship, and/or publication of this article: DESIRE (Strategies for Innovative Research to Improve Diagnosis, Prevention and Treatment in children with difficult to treat epilepsy), a FP7 funded project (Grant Agreement no: 602531), from European Commission, the grants RF-2011-02350578 and RF-2010-2319316 from Italian Ministry of Health.

## Data availability statement

The data that support the findings of this study and the codes for the analysis are available from the corresponding author upon reasonable request.

## Ethics approval statement

The study was performed according to the Declaration of Helsinki and was approved by the Ethics Committee of Fondazione IRCCS Istituto Neurologico Carlo Besta

## References

Abbasi B, Goldenholz DM. Machine learning applications in epilepsy. Epilepsia 2019; 60

Adkinson JA, Karumuri B, Hutson TN, Liu R, Alamoudi O, Vlachos I, et al. Connectivity and centrality characteristics of the epileptogenic focus using directed network analysis. IEEE Trans Neural Syst Rehabil Eng 2019; 27

Alcalá-Fdez J, Sánchez L, García S, del Jesus MJ, Ventura S, Garrell JM, et al. KEEL: A software tool to assess evolutionary algorithms for data mining problems. Soft Comput 2009; 13

Andrzejak RG, David O, Gnatkovsky V, Wendling F, Bartolomei F, Francione S, et al. Localization of Epileptogenic Zone on Pre-surgical Intracranial EEG Recordings: Toward a Validation of Quantitative Signal Analysis Approaches. Brain Topogr 2015; 28

Andrzejak RG, Schindler K, Rummel C. Nonrandomness, nonlinear dependence, and nonstationarity of electroencephalographic recordings from epilepsy patients. Phys Rev E - Stat Nonlinear, Soft Matter Phys 2012; 86

Antony AR, Alexopoulos A V., González-Martínez JA, Mosher JC, Jehi L, Burgess RC, et al. Functional connectivity estimated from intracranial EEG predicts surgical outcome in intractable temporal lobe epilepsy. PLoS One 2013; 8

Bartolomei F, Gavaret M, Hewett R, Valton L, Aubert S, Régis J, et al. Neural networks underlying parietal lobe seizures: A quantified study from intracerebral recordings. Epilepsy Res 2011; 93

Bartolomei F, Lagarde S, Wendling F, McGonigal A, Jirsa V, Guye M, et al. Defining epileptogenic networks: Contribution of SEEG and signal analysis. Epilepsia 2017; 58

Beleza P. Refractory epilepsy: A clinically oriented review. Eur Neurol 2009; 62

Bernhardt BC, Hong S, Bernasconi A, Bernasconi N. Imaging structural and functional brain networks in temporal lobe epilepsy. Front Hum Neurosci 2013

Blumenfeld H, Rivera M, Vasquez JG, Shah A, Ismail D, Enev M, et al. Neocortical and thalamic spread of amygdala kindled seizures. Epilepsia 2007; 48

Bonilha L, Jensen JH, Baker N, Breedlove J, Nesland T, Lin JJ, et al. The brain connectome as a personalized biomarker of seizure outcomes after temporal lobectomy. Neurology 2015; 84

Borgatti SP. Centrality and network flow. Soc Networks 2005; 27

Bulacio JC, Jehi L, Wong C, Gonzalez-Martinez J, Kotagal P, Nair D, et al. Long-term seizure outcome after resective surgery in patients evaluated with intracranial electrodes. Epilepsia 2012; 53

Burman RJ, Parrish RR. The widespread network effects of focal epilepsy. J Neurosci 2018; 38

Burns SP, Santaniello S, Yaffe RB, Jouny CC, Crone NE, Bergey GK, et al. Network dynamics of the brain and influence of the epileptic seizure onset zone. Proc Natl Acad Sci U S A 2014; 111

Cardinale F, Rizzi M, Vignati E, Cossu M, Castana L, d’Orio P, et al. Stereoelectroencephalography: Retrospective analysis of 742 procedures in a single centre. Brain 2019; 142

Chawla N V. Data Mining for Imbalanced Datasets: An Overview. In: Data Mining and Knowledge Discovery Handbook. 2009

Cimbalnik J, Klimes P, Sladky V, Nejedly P, Jurak P, Pail M, et al. Multi-feature localization of epileptic foci from interictal, intracranial EEG. Clin Neurophysiol 2019; 130

Coito A, Biethahn S, Tepperberg J, Carboni M, Roelcke U, Seeck M, et al. Interictal epileptogenic zone localization in patients with focal epilepsy using electric source imaging and directed functional connectivity from low-density EEG. Epilepsia Open 2019; 4

Cossu M, Cardinale F, Casaceli G, Castana L, Consales A, D’Orio P, et al. Stereo-EEG–guided radiofrequency thermocoagulations. Epilepsia 2017; 58

Cossu M, Fuschillo D, Cardinale F, Castana L, Francione S, Nobili L, et al. Stereo-EEG-guided radio-frequency thermocoagulations of epileptogenic grey-matter nodular heterotopy. J Neurol Neurosurg Psychiatry 2014; 85

David O, Blauwblomme T, Job AS, Chabards S, Hoffmann D, Minotti L, et al. Imaging the seizure onset zone with stereo-electroencephalography. Brain 2011; 134

Duncan JS, Winston GP, Koepp MJ, Ourselin S. Brain imaging in the assessment for epilepsy surgery. Lancet Neurol 2016; 15

Van Diessen E, Diederen SJH, Braun KPJ, Jansen FE, Stam CJ. Functional and structural brain networks in epilepsy: What have we learned? Epilepsia 2013; 54

Elahian B, Yeasin M, Mudigoudar B, Wheless JW, Babajani-Feremi A. Identifying seizure onset zone from electrocorticographic recordings: A machine learning approach based on phase locking value. Seizure 2017; 51

Engel J. Update on surgical treatment of the epilepsies: Summary of the second international palm desert conference on the surgical treatment of the epilepsies (1992). Neurology 1993; 43

Englot DJ, Hinkley LB, Kort NS, Imber BS, Mizuiri D, Honma SM, et al. Global and regional functional connectivity maps of neural oscillations in focal epilepsy. Brain 2015; 138

Englot DJ, Konrad PE, Morgan VL. Regional and global connectivity disturbances in focal epilepsy, related neurocognitive sequelae, and potential mechanistic underpinnings. Epilepsia 2016; 57

Englot DJ, Morgan VL, Chang C. Impaired vigilance networks in temporal lobe epilepsy: Mechanisms and clinical implications. Epilepsia 2020; 61

Fiest KM, Sauro KM, Wiebe S, Patten SB, Kwon CS, Dykeman J, et al. Prevalence and incidence of epilepsy. Neurology 2017; 88

Fornito A, Zalesky A, Bullmore ET. Fundamentals of Brain Network Analysis. 2016

Frauscher B. Localizing the epileptogenic zone. Curr Opin Neurol 2020; 33

Di Giacomo R, Uribe-San-Martin R, Mai R, Francione S, Nobili L, Sartori I, et al. Stereo-EEG ictal/interictal patterns and underlying pathologies. Seizure 2019; 72

Gnatkovsky V, De Curtis M, Pastori C, Cardinale F, Lo Russo G, Mai R, et al. Biomarkers of epileptogenic zone defined by quantified stereo-EEG analysis. Epilepsia 2014; 55

Gnatkovsky V, Pelliccia V, de Curtis M, Tassi L. Two main focal seizure patterns revealed by intracerebral electroencephalographic biomarker analysis. Epilepsia 2019; 60

Grobelny BT, London D, Hill TC, North E, Dugan P, Doyle WK. Betweenness centrality of intracranial electroencephalography networks and surgical epilepsy outcome. Clin Neurophysiol. 2018 Sep;129(9):1804–1812. doi: 10.1016/j.clinph.2018.02.135. Epub 2018 Mar 19. PMID: 29981955.

He X, Doucet GE, Pustina D, Sperling MR, Sharan AD, Tracy JI. Presurgical thalamic ‘hubness’ predicts surgical outcome in temporal lobe epilepsy. Neurology 2017; 88

Khalilov I, Holmes GL, Ben-Ari Y. In vitro formation of a secondary epileptogenic mirror focus by interhippocampal propagation of seizures. Nat Neurosci 2003; 6

Khambhati AN, Bassett DS, Oommen BS, Chen SH, Lucas TH, Davis KA, et al. Recurring functional interactions predict network architecture of interictal and ictal states in neocortical epilepsy. eNeuro 2017; 4

Kuncheva LI. Combining pattern classifiers: methods and algorithms. New York: Wiley; 2004

Lagarde S, Roehri N, Lambert I, Trebuchon A, McGonigal A, Carron R, et al. Interictal stereotactic-EEG functional connectivity in refractory focal epilepsies. Brain 2018; 141

Lagarde S, Bénar CG, Wendling F, Bartolomei F. Interictal Functional Connectivity in Focal Refractory Epilepsies Investigated by Intracranial EEG. Brain Connect. 2022 Dec;12(10):850–869. doi: 10.1089/brain.2021.0190. Epub 2022 Sep 14. PMID: 35972755; PMCID: PMC9807250.

Lehnertz K, Bialonski S, Horstmann MT, Krug D, Rothkegel A, Staniek M, et al. Synchronization phenomena in human epileptic brain networks. J Neurosci Methods 2009; 183

López V, Fernández A, García S, Palade V, Herrera F. An insight into classification with imbalanced data: Empirical results and current trends on using data intrinsic characteristics. Inf Sci (Ny) 2013; 250

Lüders HO, Najm I, Nair D, Widdess-Walsh P, Bingman W. The epileptogenic zone: General principles. In: Epileptic Disorders. 2006

Marino AC, Yang GJ, Tyrtova E, Wu K, Zaveri HP, Farooque P, et al. Resting state connectivity in neocortical epilepsy: The epilepsy network as a patient-specific biomarker. Clin Neurophysiol 2019; 130

Moseley BD, Sinha S, Meyer FB, Marsh WR, Britton JW. Long term outcomes in patients with preoperative generalized interictal epileptiform abnormalities following amygdalohippocampectomy. Epilepsy Res 2012; 99

Oldham S, Fulcher B, Parkes L, Arnatkeviciūtė A, Suo C, Fornito A. Consistency and differences between centrality measures across distinct classes of networks. PLoS One 2019; 14

Panzica F, Varotto G, Rotondi F, Spreafico R, Franceschetti S. Identification of the epileptogenic zone from stereo-EEG signals: A connectivity-graph theory approach. Front Neurol 2013; 4 NOV

Paulo DL, Wills KE, Johnson GW, Gonzalez HFJ, Rolston JD, Naftel RP, Reddy SB, Morgan VL, Kang H, Williams Roberson S, Narasimhan S, Englot DJ. SEEG Functional Connectivity Measures to Identify Epileptogenic Zones: Stability, Medication Influence, and Recording Condition. Neurology. 2022 May 17;98(20):e2060–e2072. doi: 10.1212/WNL.0000000000200386. Epub 2022 Mar 25. PMID: 35338075; PMCID: PMC9162047.

Pijn JP, Lopes da Silva F. Propagation of Electrical Activity: Nonlinear Associations and Time Delays between EEG Signals. In: Basic Mechanisms of the EEG. 1993

Proix T, Bartolomei F, Chauvel P, Bernard C, Jirsa VK. Permittivity coupling across brain regions determines seizure recruitment in partial epilepsy. J Neurosci 2014; 34

Rossi Sebastiano D, Tassi L, Duran D, Visani E, Gozzo F, Cardinale F, et al. Identifying the epileptogenic zone by four non-invasive imaging techniques versus stereo-EEG in MRI-negative pre-surgery epilepsy patients. Clin Neurophysiol 2020; 131

Ryvlin P, Cross JH, Rheims S. Epilepsy surgery in children and adults. Lancet Neurol 2014; 13

Sabesan S, Good LB, Tsakalis KS, Spanias A, Treiman DM, Iasemidis LD. Information flow and application to epileptogenic focus localization from intracranial EEG. IEEE Trans Neural Syst Rehabil Eng 2009; 17

Sinha N, Johnson GW, Davis KA, Englot DJ. Integrating Network Neuroscience Into Epilepsy Care: Progress, Barriers, and Next Steps. Epilepsy Curr. 2022 May 13;22(5):272–278. doi: 10.1177/15357597221101271. PMID: 36285209; PMCID: PMC9549227.

Spencer S, Huh L. Outcomes of epilepsy surgery in adults and children. Lancet Neurol 2008; 7

Sun S, Zhang C. Subspace ensembles for classification. Phys A Stat Mech its Appl 2007; 385

Tang S, Chen SP. The generation mechanism of synthetic minority class examples. In: 5th Int. Conference on Information Technology and Applications in Biomedicine, ITAB 2008 in conjunction with 2nd Int. Symposium and Summer School on Biomedical and Health Engineering, IS3BHE 2008. 2008

Tomlinson SB, Porter BE, Marsh ED. Interictal network synchrony and local heterogeneity predict epilepsy surgery outcome among pediatric patients. Epilepsia 2017; 58

Traub RD, Whittington MA, Buhl EH, LeBeau FEN, Bibbig A, Boyd S, et al. A possible role for gap junctions in generation of very fast EEG oscillations preceding the onset of, and perhaps initiating, seizures. Epilepsia 2001; 42

Ung H, Cazares C, Nanivadekar A, Kini L, Wagenaar J, Becker D, et al. Interictal epileptiform activity outside the seizure onset zone impacts cognition. Brain 2017; 140

Varotto G, Tassi L, Franceschetti S, Spreafico R, Panzica F. Epileptogenic networks of type II focal cortical dysplasia: A stereo-EEG study. Neuroimage 2012; 61

Varotto G, Susi G, Tassi L, Gozzo F, Franceschetti S, Panzica F. Comparison of Resampling Techniques for Imbalanced Datasets in Machine Learning: Application to Epileptogenic Zone Localization From Interictal Intracranial EEG Recordings in Patients With Focal Epilepsy. Front Neuroinform. 2021 Nov 19;15:715421. doi: 10.3389/fninf.2021.715421. PMID: 34867255; PMCID: PMC8641296.

Vaugier L, Lagarde S, McGonigal A, Trébuchon A, Milh M, Lépine A, et al. The role of stereoelectroencephalography (SEEG) in reevaluation of epilepsy surgery failures. Epilepsy Behav 2018; 81

Wendling F, Chauvel P, Biraben A, Bartolomei F. From intracerebral EEG signals to brain connectivity:Identification of epileptogenic networks in partial epilepsy. Front Syst Neurosci 2010; 4

Wilke C, Worrell G, He B. Graph analysis of epileptogenic networks in human partial epilepsy. Epilepsia 2011; 52

Wu T, Ge S, Zhang R, Liu H, Chen Q, Zhao R, et al. Neuromagnetic coherence of epileptic activity: An MEG study. Seizure 2014; 23

Yuan J, Chen Y, Hirsch E. Intracranial electrodes in the presurgical evaluation of epilepsy. Neurol Sci 2012; 33

