## Supplementary materials for "Interictal epileptic network hubs as a biomarker for automatic localization of the epileptogenic zone: a connectivity and machine learning based analysis of stereo-EEG"

**Supplementary Table 1. Description of the set of graph-theory based centrality measures used in this study.**

| **Centrality index** | **Expression** | **Description** |
| --- | --- | --- |
| Outdegree c. | $C_{OD}\left( i \right)= \sum_{j\neq i} sgn (w_{ij})$  where $sgn(x)$ is the sign function. | Number of edges outcoming from the considered node |
| Indegree c. | $C_{ID}\left( i \right)= \sum_{j\neq i} {sgn (w}_{ji})$ | Number of edges incoming to the considered node |
| Outstrength c. | $C_{OS}\left( i \right)= \sum_{j\neq i} w_{ij}$ | Sum of the edge weights outcoming from the considered node |
| Instrength c. | $C_{IS}\left( i \right)= \sum_{j\neq i} w_{ji}$ | Sum of the edge weights incoming to the considered node |
| Betweenness c. | $C_{B}\left( i \right)= \frac{1}{(N-1) (N-2)} \sum_{\begin{aligned} h\neq i \\ h\neq j \\ j\neq i \end{aligned}} \frac{\rho_{hj}(i)}{\rho_{hj}}$  where $\rho_{hj}(i)$ is the number of shortest paths between j and k that pass through *i*, $\rho_{hj}$ is the number of shortest paths between *j* and *k*, and $(N-1)(N-2)$ is the number of node pairs that does not include node *i*. | Measures of how often the considered node appears on a shortest path between two nodes in the graph. |
| Outcloseness c. | $C_{OC}\left( i \right)=\frac{1}{\sum_{j=1}^{N} d_{ij}}$  Where $d_{ij}$ is the topological distance from node *i* to node *j* | It measures the inverse sum of the distance of the shortest paths from the considered node to all other nodes in the graph |
| Incloseness c. | $C_{IC}\left( i \right)=\frac{1}{\sum_{j=1}^{N} d_{ji}}$  Where $d_{ji}$ is the distance from all the nodes to the node *i*. | It measures the inverse sum of the distance of the shortest paths from all nodes of the graph to the considered node. |
| Page Rank c. | $C_{PR}\left( i \right)={D (D-\alpha A)}^{-1}1$,  where 1 is a column vector of ones of length *N_L_* , *D* is the diagonal matrix of node outdegrees, and *α* is a free parameter which weights the contribution of network topology to the centrality score. | This measure scales the contributions that the neighbors of node *i* make to its centrality by the degree of those neighbors, thereby accounting for any potential bias associated with links to highly connected nodes. |
| Eigenvector c. | $C_{E} (i) =\frac{1}{\lambda_{1}}\sum_{j=1}^{N} A_{ij}C_{E}(j)$  Where A is the adjacency matrix and *λ_1_* its largest eigenvalue. | Recursive measure that considers both the degree of the considered node and the degree of its neighbors. |
